# Longitudinal associations of epigenetic aging with cognitive aging in Hispanic/Latino adults from the Hispanic Community Health Study/Study of Latinos

**DOI:** 10.1101/2025.04.03.25325181

**Authors:** Myriam Fornage, Wassim Tarraf, Rui Xia, Adriana Ordonez, Tamar Sofer, Freddie Márquez, Bharat Thyagarajan, Gregory A Talavera, Linda C. Gallo, Charles DeCarli, Hector M. González

## Abstract

Due to the paucity of longitudinal DNA methylation data (DNAm), especially among Hispanic/Latino adults, the association between changes in epigenetic clocks over time and cognitive aging phenotypes has not been investigated. This longitudinal study included 2671 Hispanic/Latino adults (57 years; 66% women) with blood DNAm data and neurocognitive function assessed at two visits approximately 7 years apart. We evaluated the associations of 5 epigenetic clocks and their between-visit change with multiple measures of cognitive aging that included a global cognitive function score at each visit, between-visit change in global cognitive function score, MCI diagnosis, and presence of significant cognitive decline at visit 2 (V2). There were significant associations between greater acceleration for all clocks and lower global cognitive function at each visit. The strongest associations were observed for GrimAge and DunedinPACE. Similar results were observed for domain-specific cognitive function at each visit and MCI diagnosis at V2. There was a significant association of decline in global cognitive function with increase in age acceleration between the two visits for PhenoAge and GrimAge. Between-visit increase in age acceleration for these two clocks was also associated with a greater risk of MCI diagnosis and presence of significant cognitive decline at V2. Epigenetic aging is associated with lower global and domain-specific cognitive function, greater cognitive decline, and greater risk of MCI in Hispanic/Latino adults. Longitudinal assessment of change in age acceleration for second-generation clocks, GrimAge and PhenoAge may provide additional value in predicting cognitive aging beyond a single time point assessment.

## Introduction

Aging can be defined as the time-related deterioration of physiological function that leads to physical and functional decline and vulnerability to chronic conditions, such as cognitive decline and dementia ^1^. There is considerable interindividual variation in the rate of aging, and individuals with the same chronological age can differ in their level of age-dependent biological changes, known as biological age. The development of biomarkers of biological age, which can be used to assess the effectiveness of aging interventions and accurately predict age-related conditions and mortality, is an expanding area of research ^2^.

DNA methylation (DNAm) is an epigenetic mechanism by which genes and environmental, lifestyle, and sociocultural exposures dynamically interact to regulate gene expression, thereby shaping various traits related to health and aging. Most human tissues and cell types exhibit profound changes in DNAm patterns with advancing age ^3,4^, and multiple DNAm signatures of aging have been characterized across various human tissues ^5–11^. These “epigenetic clocks” are thought to capture different aspects of the multidimensional aging process. While first-generation epigenetic clocks were developed to specifically predict chronological age ^6,7,11^, second- and third-generation epigenetic clocks have been recently proposed to predict biological age represented by various clinical biomarkers of physiological dysregulation and change in health indicators ^5,8–10^. Age acceleration, the deviation of the DNAm-estimated age from the chronological age, has been proposed as a novel biomarker of aging. Indeed, positive age acceleration, where a person’s biological age is older than their chronological age, has been associated with a greater risk of various diseases and mortality ^12^.

Due to the paucity of longitudinal DNAm data, especially among Hispanic/Latino adults, the association between changes in age acceleration over time and cognitive aging phenotypes has not been widely investigated.

We leveraged data from the Study of Latinos-Investigation of Neurocognitive Aging (SOL-INCA) to examine the associations of three generation epigenetic clocks and their change over a 7-year period, with cognitive decline and mild cognitive impairment (MCI) in Hispanic/Latino middle-aged and older adults.

## Results

Characteristics of the study sample are shown in Table 1. The mean age of the at visit 1 was 57 years and there were 66% women. On average, educational attainment was 11 years. Approximately, one third of the study sample had an ideal cardiovascular heath as measured by American Heart Association (AHA) Life’s Simple 7, while 29% had poor cardiovascular health. The frequency of MCI in the sample was 21% and that of significant cognitive decline was 50%. Estimated DNA methylation age by each of the 5 clocks was strongly correlated with chronological age at each visit (r^2^ from 0.76 to 0.86, Supplementary Table 1), with the strongest correlations observed for GrimAge. For each clock, measures of age acceleration were strongly correlated across the two visits (r^2^ from 0.81 to 0.93, Supplementary Table 2). At visit 1, the correlations among measures of age acceleration by the various clocks varied between 0.18 to 0.88, with the strongest correlation observed between the Horvath and Hannum clocks. Similar correlations were observed at visit 2 (Supplementary Table 2).

**Table 1.**
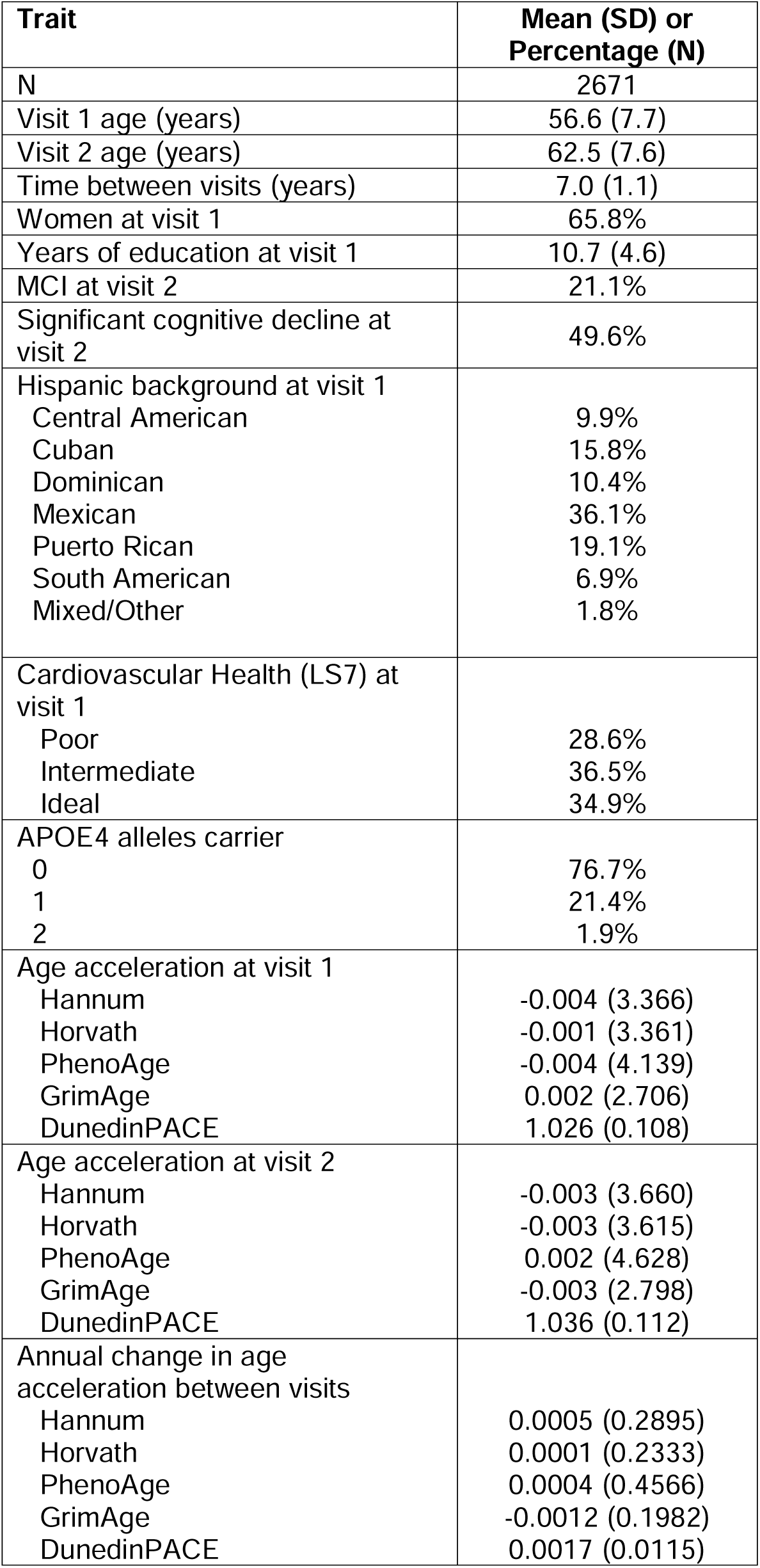
Descriptive characteristics of the study participants.

### Cross-sectional associations of epigenetic clocks with cognitive aging measures

Cross-sectional associations of the five epigenetic clocks with the global cognitive function score are shown at each visit in Table 2. There were significant associations between greater age acceleration for all clocks and lower global cognitive function at both visits. The strongest associations were observed for GrimAge and DunedinPACE. Adjustments for education, language preference, and cardiovascular health did not meaningfully attenuate these associations. Cross-sectional associations of the five epigenetic clocks with individual cognitive tests showed similar results (Supplementary Table 3), although there was no association of the first-generation clocks with B-SEVLT sum or B-SEVLT recall. Associations of second- and third-generation clocks were strongest with WF and DSST.

**Table 2.** Cross-sectional associations of global cognitive function at visit 1 and at visit 2 with epigenetic age acceleration (EAA) for 5 clocks at the same visit.

|  |  | Model 1 |  |  | Model 2 |  |  | Model 3 |  |  |
| --- | --- | --- | --- | --- | --- | --- | --- | --- | --- | --- |
|  | **EAA Measure** | beta | SE | P value | beta | SE | P value | beta | SE | P value |
| Visit 1 | Hannum | -0.009 | 0.003 | **$4.4 \times 10^{-3}$** | -0.007 | 0.003 | **$1.6 \times 10^{-2}$** | -0.007 | 0.003 | **$2.1 \times 10^{-2}$** |
|  | Horvath | -0.011 | 0.003 | **$8.3 \times 10^{-4}$** | -0.008 | 0.003 | **$2.9 \times 10^{-3}$** | -0.008 | 0.003 | **$4.6 \times 10^{-3}$** |
|  | PhenoAge | -0.011 | 0.003 | **$1.7 \times 10^{-5}$** | -0.008 | 0.002 | **$6.8 \times 10^{-4}$** | -0.007 | 0.002 | **$2.2 \times 10^{-3}$** |
|  | GrimAge | -0.018 | 0.004 | **$9.1 \times 10^{-6}$** | -0.012 | 0.004 | **$8.0 \times 10^{-4}$** | -0.011 | 0.004 | **$3.8 \times 10^{-3}$** |
|  | DunedinPACE * | -0.011 | 0.002 | **$6.5 \times 10^{-10}$** | -0.008 | 0.002 | **$9.6 \times 10^{-7}$** | -0.007 | 0.002 | **$1.3 \times 10^{-5}$** |
| Visit 2 | Hannum | -0.013 | 0.004 | **$9.0 \times 10^{-4}$** | -0.010 | 0.003 | **$2.7 \times 10^{-3}$** | -0.009 | 0.003 | **$5.7 \times 10^{-3}$** |
|  | Horvath | -0.013 | 0.004 | **$8.0 \times 10^{-4}$** | -0.010 | 0.003 | **$2.4 \times 10^{-3}$** | -0.010 | 0.003 | **$3.9 \times 10^{-3}$** |
|  | PhenoAge | -0.016 | 0.003 | **$9.5 \times 10^{-8}$** | -0.014 | 0.003 | **$1.6 \times 10^{-7}$** | -0.011 | 0.002 | **$3.3 \times 10^{-6}$** |
|  | GrimAge | -0.032 | 0.005 | **$3.3 \times 10^{-10}$** | -0.026 | 0.004 | **$6.3 \times 10^{-9}$** | -0.024 | 0.005 | **$2.7 \times 10^{-7}$** |
|  | DunedinPACE * | -0.013 | 0.002 | **$2.2 \times 10^{-10}$** | -0.009 | 0.002 | **$1.7 \times 10^{-7}$** | -0.008 | 0.002 | **$1.6 \times 10^{-5}$** |

Model 1: Adjusted for age, gender, center, and Hispanic background; Model 2: Adjusted for variables in Model 1 + years of education, language preference; Model 3: Adjusted for variables in Model 2 + cardiovascular health score (Life's Simple 7 category); \* DunedinPACE was rescaled to allow for comparison with other clocks (P-value shown for association on the original scale)

Associations of MCI status at visit 2 with epigenetic clocks measured at each visit are shown in Table 3. The strongest associations were observed for second- and third-generation clocks, GrimAge, and DunedinPACE at either visit, with an estimated 3 to 9% increase in risk of MCI. Associations of first-generation clocks were generally weaker, especially for visit 1 data. Further adjustment for education, language preference, and cardiovascular health mildly attenuated association results for all clocks but, except for Hannum and PhenoAge measured at visit 1, associations remained significant.

**Table 3.** Association of MCI status at visit 2 with age acceleration (EAA) for 5 clocks at visit 1 (V1) and visit 2 (V2).

|  | Model 1 |  |  | Model 2 |  |  | Model 3 |  |  |
| --- | --- | --- | --- | --- | --- | --- | --- | --- | --- |
| **EAA Measure** | OR | 95% CI | P value | OR | 95% CI | P value | OR | 95% CI | P value |
| V1 Hannum | 1.03 | 1.00; 1.06 | **$1.6 \times 10^{-2}$** | 1.03 | 1.00; 1.06 | **$2.4 \times 10^{-2}$** | 1.03 | 1.00; 1.06 | $5.3 \times 10^{-2}$ |
| V1 Horvath | 1.04 | 1.01; 1.07 | **$7.2 \times 10^{-3}$** | 1.04 | 1.01; 1.07 | **$1.1 \times 10^{-2}$** | 1.03 | 1.00; 1.06 | **$2.6 \times 10^{-2}$** |
| V1 PhenoAge | 1.03 | 1.01; 1.06 | **$5.7 \times 10^{-3}$** | 1.03 | 1.01; 1.05 | **$1.3 \times 10^{-2}$** | 1.02 | 1.00; 1.05 | $7.5 \times 10^{-2}$ |
| V1 GrimAge | 1.07 | 1.04; 1.11 | **$5.4 \times 10^{-5}$** | 1.07 | 1.03; 1.11 | **$1.0 \times 10^{-4}$** | 1.06 | 1.02; 1.10 | **$3.5 \times 10^{-3}$** |
| V1 DunedinPACE * | 1.04 | 1.03; 1.06 | **$1.5 \times 10^{-7}$** | 1.04 | 1.02; 1.05 | **$9.2 \times 10^{-7}$** | 1.03 | 1.02; 1.05 | **$1.0 \times 10^{-4}$** |
| V2 Hannum | 1.04 | 1.01; 1.07 | **$4.3 \times 10^{-3}$** | 1.04 | 1.01; 1.06 | **$5.7 \times 10^{-3}$** | 1.03 | 1.00; 1.06 | **$1.9 \times 10^{-2}$** |
| V2 Horvath | 1.04 | 1.02; 1.07 | **$1.0 \times 10^{-3}$** | 1.04 | 1.02; 1.07 | **$1.5 \times 10^{-3}$** | 1.04 | 1.01; 1.07 | **$4.6 \times 10^{-3}$** |
| V2 PhenoAge | 1.04 | 1.02; 1.05 | **$2.0 \times 10^{-4}$** | 1.04 | 1.02; 1.05 | **$3.0 \times 10^{-4}$** | 1.03 | 1.01; 1.05 | **$4.7 \times 10^{-3}$** |
| V2 GrimAge | 1.09 | 1.05; 1.12 | **$1.4 \times 10^{-6}$** | 1.09 | 1.05; 1.13 | **$1.4 \times 10^{-6}$** | 1.07 | 1.03; 1.11 | **$1.2 \times 10^{-4}$** |
| V2 DunedinPACE * | 1.03 | 1.02; 1.05 | **$2.3 \times 10^{-6}$** | 1.03 | 1.02; 1.05 | **$7.6 \times 10^{-6}$** | 1.02 | 1.01; 1.04 | **$5 \times 10^{-4}$** |

Model 1: Adjusted for age, gender, center, and Hispanic background; Model 2: Adjusted for variables in Model 1 + years of education, language preference; Model 3: Adjusted for variables in Model 2 + cardiovascular health (Life's Simple 7 category); \* DunedinPACE was rescaled to allow for comparison with other clocks (P-value shown for association on the original scale)

Associations of epigenetic clocks with the presence of significant cognitive decline at visit 2 are shown in Table 4. There were significant associations of second- and third-generation clocks PhenoAge, GrimAge, and DunedinPACE measured at visit 2 with presence of significant cognitive decline (Odds Ratio (OR)=1.02 to 1.07, P=7.1x10^-6^ to 9.0x10^-3^). When epigenetic clocks were measured at visit 1, only GrimAge was significantly associated with presence of significant cognitive decline (OR=1.05, P=1.6x10^-3^). These associations remained significant with further adjustment for education, language preference, and cardiovascular health. When cognitive change between the two visits was modeled as a quantitative trait, greater age acceleration measured at visit 1 or visit 2 was associated with decline in global cognitive function between visits for all clocks, although associations were weaker or not statistically significant for first-generation clocks (Supplementary Table 4).

**Table 4.** Association of presence of significant cognitive decline at visit 2 with epigenetic age acceleration (EAA) for 5 clocks at visit 1 and visit 2.

|  | Model 1 |  |  | Model 2 |  |  | Model 3 |  |  |
| --- | --- | --- | --- | --- | --- | --- | --- | --- | --- |
| **EAA Measure** | OR | 95% CI | P value | OR | 95% CI | P value | OR | 95% CI | P value |
| V1 Hannum | 1.01 | 0.99; 1.03 | 0.29 | 1.01 | 0.99; 1.04 | 0.28 | 1.01 | 0.99; 1.04 | 0.32 |
| V1 Horvath | 1.01 | 0.98; 1.03 | 0.59 | 1.01 | 0.98; 1.03 | 0.57 | 1.01 | 0.98; 1.03 | 0.63 |
| V1 PhenoAge | 1.01 | 0.99; 1.03 | 0.27 | 1.01 | 0.99; 1.03 | 0.24 | 1.01 | 0.99; 1.03 | 0.37 |
| V1 GrimAge | 1.05 | 1.02; 1.08 | **$1.6 \times 10^{-3}$** | 1.05 | 1.02; 1.08 | **$9.0 \times 10^{-4}$** | 1.04 | 1.02; 1.08 | **$3.0 \times 10^{-3}$** |
| V1 DunedinPACE * | 1.01 | 1.00; 1.02 | 0.06 | 1.01 | 1.00; 1.03 | **$4.4 \times 10^{-2}$** | 1.01 | 1.00; 1.02 | 0.10 |
| V2 Hannum | 1.01 | 0.99; 1.03 | 0.32 | 1.01 | 0.99; 1.03 | 0.29 | 1.01 | 0.99; 1.03 | 0.37 |
| V2 Horvath | 1.01 | 0.99; 1.03 | 0.44 | 1.01 | 0.99; 1.03 | 0.42 | 1.01 | 0.99; 1.03 | 0.50 |
| V2 PhenoAge | 1.02 | 1.01; 1.04 | **$9.0 \times 10^{-3}$** | 1.02 | 1.01; 1.04 | **$7.0 \times 10^{-3}$** | 1.02 | 1.00; 1.04 | **$2.0 \times 10^{-2}$** |
| V2 GrimAge | 1.07 | 1.04; 1.10 | **$7.1 \times 10^{-6}$** | 1.07 | 1.04; 1.10 | **$2.8 \times 10^{-6}$** | 1.07 | 1.04; 1.10 | **$1.2 \times 10^{-5}$** |
| V2 DunedinPACE * | 1.02 | 1.01; 1.03 | **$8.0 \times 10^{-4}$** | 1.02 | 1.01; 1.03 | **$5.0 \times 10^{-4}$** | 1.02 | 1.01; 1.03 | **$1.5 \times 10^{-3}$** |

Model 1: Adjusted for age, gender, center, and Hispanic background; Model 2: Adjusted for variables in Model 1 + years of education, language preference; Model 3: Adjusted for variables in Model 2 + cardiovascular health (Life's Simple 7 category); \* DunedinPACE was rescaled to allow for comparison with other clocks (P-value shown for association on the original scale)

### Longitudinal associations of epigenetic clocks with cognitive aging measures

We next examined the association of change in global cognitive function with change in epigenetic clocks between the two visits (Table 5). Controlling for visit 1 PhenoAge acceleration, an increase in PhenoAge acceleration from visit 1 to visit 2 was associated with a decline in global cognitive function between the two visits (P=0.021). Similarly, an increase in GrimAge acceleration, controlling for visit 1 GrimAge acceleration, was associated with a decline in global cognitive function between the two visits (P=0.016). Further adjustment for education, language preference, and cardiovascular health did not meaningfully change these results. Association of change in individual cognitive test scores with change in epigenetic clocks between the two visits showed varying results (Supplementary Table 5). Change in PhenoAge acceleration was associated with a decline in B-SEVLT sum and DSST (P=0.048 and 0.0006, respectively), while change in GrimAge acceleration was associated with a decline in WF and DSST (P=0.015 and 0.0004, respectively). Interestingly, change in DunedinPACE between visits was also associated with decline in DSST, although adjusting for cardiovascular health mitigated this relationship.

**Table 5.** Association of global cognitive function change with change (Δ) in epigenetic age acceleration (EAA) between visit 1 and visit 2 for 5 clocks.

|  | Model 1 |  |  | Model 2 |  |  | Model 3 |  |  |
| --- | --- | --- | --- | --- | --- | --- | --- | --- | --- |
| **EAA Measure** | beta | SE | P value | beta | SE | P value | beta | SE | P value |
| $\Delta$ Hannum | 0.024 | 0.067 | 0.718 | 0.025 | 0.067 | 0.711 | 0.035 | 0.068 | 0.605 |
| $\Delta$ Horvath | -0.065 | 0.083 | 0.436 | -0.066 | 0.083 | 0.427 | -0.059 | 0.083 | 0.477 |
| $\Delta$ PhenoAge | -0.099 | 0.043 | **0.021** | -0.106 | 0.042 | **0.013** | -0.097 | 0.043 | **0.024** |
| $\Delta$ GrimAge | -0.239 | 0.099 | **0.016** | -0.256 | 0.099 | **0.009** | -0.238 | 0.099 | **0.016** |
| $\Delta$ DunedinPACE * | -0.043 | 0.028 | 0.121 | -0.043 | 0.028 | 0.125 | -0.037 | 0.028 | 0.187 |

Model 1: Adjusted for V1 age acceleration, age, gender, center, and Hispanic background; Model 2: Adjusted for variables in Model 1 + years of education, language preference; Model 3: Adjusted for variables in Model 2 + cardiovascular health (Life's Simple 7 category); \* DunedinPACE was rescaled to allow for comparison with other clocks (P-value shown for association on the original scale)

Tables 6 presents the association of change in epigenetic clocks between the two visits with presence of MCI and significant cognitive decline at visit 2. Increase in age acceleration between the two visits for PhenoAge, and GrimAge was associated with a greater risk of MCI, controlling for visit 1 age acceleration values (OR=1.3, and 1.8; P=0.017 and 0.010, respectively). Similar findings were observed for association between presence of significant cognitive decline and change in PhenoAge and GrimAge acceleration (OR=1.3 and 2.2; P=0.0025 and 0.0001, respectively). Between visit change in DunedinPACE was associated with a 20% greater risk of significant cognitive decline (P=0.0014). Further adjustment for education, language preference, and cardiovascular health did not meaningfully change these results.

**Table 6.** Association of presence of MCI and significant cognitive decline at visit 2 with change (Δ) in epigenetic age acceleration (EAA) between visit 1 and visit 2 for 5 clocks.

|  |  | Model 1 |  |  | Model 2 |  |  | Model 3 |  |  |
| --- | --- | --- | --- | --- | --- | --- | --- | --- | --- | --- |
|  | **EAA Measure** | OR | 95% CI | P value | OR | 95% CI | P value | OR | 95% CI | P value |
| MCI | $\Delta$ Hannum | 1.23 | 0.89; 1.69 | 0.21 | 1.23 | 0.89; 1.71 | 0.21 | 1.17 | 0.85; 1.63 | 0.34 |
| | $\Delta$ Horvath | 1.46 | 1.00; 2.16 | 0.05 | 1.48 | 1.00; 2.19 | 0.05 | 1.43 | 1.03; 2.11 | 0.08 |
| | $\Delta$ PhenoAge | 1.28 | 1.04; 1.57 | **$1.7 \times 10^{-2}$** | 1.32 | 1.07; 1.62 | **$9.0 \times 10^{-3}$** | 1.26 | 1.02; 1.55 | **$3.3 \times 10^{-2}$** |
| | $\Delta$ GrimAge | 1.85 | 1.16; 2.98 | **$1.0 \times 10^{-2}$** | 2.00 | 1.24; 3.24 | **$4.0 \times 10^{-3}$** | 1.86 | 1.15; 3.02 | **$1.2 \times 10^{-2}$** |
| | $\Delta$ DunedinPACE * | 1.07 | 0.94; 1.22 | 0.32 | 1.07 | 0.94; 1.23 | 0.29 | 1.04 | 0.91; 1.20 | 0.53 |
| Significant Cognitive Decline | $\Delta$ Hannum | 1.03 | 0.79; 1.35 | 0.80 | 1.04 | 0.80; 1.36 | 0.75 | 1.02 | 0.78; 1.33 | 0.88 |
| | $\Delta$ Horvath | 1.11 | 0.80; 1.54 | 0.54 | 1.12 | 0.80; 1.55 | 0.51 | 1.09 | 0.78; 1.52 | 0.61 |
| | $\Delta$ PhenoAge | 1.30 | 1.09; 1.55 | **$2.5 \times 10^{-3}$** | 1.31 | 1.10; 1.56 | **$2.2 \times 10^{-3}$** | 1.28 | 1.08; 1.52 | **$5.0 \times 10^{-3}$** |
| | $\Delta$ GrimAge | 2.19 | 1.47; 3.26 | **$1.0 \times 10^{-4}$** | 2.23 | 1.49; 3.33 | **$8.1 \times 10^{-5}$** | 2.20 | 1.47; 3.29 | **$1.1 \times 10^{-4}$** |
| | $\Delta$ DunedinPACE * | 1.20 | 1.07; 1.34 | **$1.4 \times 10^{-3}$** | 1.20 | 1.08; 1.35 | **$1.0 \times 10^{-3}$** | 1.20 | 1.07; 1.34 | **$1.6 \times 10^{-3}$** |

Model 1: Adjusted for V1 age acceleration, age, gender, center, and Hispanic background; Model 2: Adjusted for variables in Model 1 + years of education, language preference; Model 3: Adjusted for variables in Model 2 + cardiovascular health (Life's Simple 7 category); \* DunedinPACE was rescaled to allow for comparison with other clocks (P-value shown for association on the original scale)

### Comparison with APOE4 effects

We sought to compare the magnitude of effects of epigenetic clocks on measures of cognitive aging to the magnitude of effects of APOE4 on the same cognitive measures. However, in this sample, there was no association between APOE4 and measures of cognitive aging (not shown), except for between-visit change in DSST. The magnitude of effects of V1 second- and third-generation clocks PhenoAge, GrimAge, and DunedinPACE on between-visit decline in DSST was considerably smaller (3-to 8-fold) than that of APOE4 (Supplementary Table 6). In contrast, the magnitude of effects of between-visit change in age acceleration on decline in DSST was similar to that of APOE4 for PhenoAge and DunedinPACE, while it was almost 4-fold larger for GrimAge (Supplementary Table 6).

### Association of cognitive aging measures with surrogate biomarker components of GrimAge

We also evaluated the association of cognitive aging measures with acceleration in a new version of GrimAge (GrimAge2^13^) and with its 10 surrogate biomarker components. Acceleration in PC-based GrimAge, GrimAge (original version), and GrimAge2 performed similarly in association analyses of the various cognitive aging measures (Supplementary Table 7). Estimated effect sizes were slightly larger for PC-based GrimAge while strength of associations was slightly larger for GrimAge2, especially for V2 data. In addition, there were strong associations of several surrogate biomarker components of GrimAge2 with each of the cognitive aging measures (Supplementary Table 8). DNAm surrogate of hemoglobin A1C exhibited the strongest associations across cognitive aging measures and visits.

## Discussion

Our study is the first to investigate the longitudinal association of epigenetic aging with cognitive aging in a large sample of Hispanic/Latino adults. We compared the association of five epigenetic clocks, including first-generation clocks Hannum and Horvath, second-generation clocks PhenoAge and GrimAge, and third-generation clock DunedinPACE, with multiple measures of cognitive aging. In line with published data ^5,14,15^, the five epigenetic clocks showed significant inter-correlations, with clocks from the same generation being more strongly correlated with one another and clocks from more distant generations being less strongly correlated. These results are consistent with the observation that different generation epigenetic clocks capture distinct aspects of the aging process. First-generation epigenetic clocks were developed to predict chronological age ^6,7^. Second-generation epigenetic clocks were developed to predict mortality using various physiological biomarkers ^8,9^. Third-generation clock DunedinPACE was developed to predict change in health indicators from early adulthood to middle age ^5^.

In cross-sectional analyses, greater age acceleration of all clocks was associated with lower general and domain-specific cognitive function at each visit and with greater risk of MCI at visit 2, while only second- and third-generation clocks were associated with presence of significant cognitive decline at visit 2. Consistently, in longitudinal analyses, increase in second-generation clocks PhenoAge acceleration and GrimAge acceleration was associated with decline in global and domain-specific cognitive function between visits. Across all analyses, GrimAge acceleration and between-visit change in GrimAge acceleration had generally the strongest estimated association with cognitive aging, adding to the growing literature suggesting that accelerated GrimAge represents a valuable biomarker of declining brain health ^16,17^. GrimAge is a composite biomarker of blood DNAm surrogates of smoking pack-years and 7 plasma proteins, including adrenomedullin, β2-microglobulin, growth differentiation factor 15, plasminogen activator inhibitor 1 and tissue inhibitor metalloproteinase 1. The new version of GrimAge, GrimAge2, additionally includes DNAm surrogates of hemoglobin A1C and C-reactive protein, and has been shown to outperform GrimAge in predicting mortality and age-related health conditions ^13^. All these plasma proteins and smoking have been associated with cognitive impairment, cognitive decline, and dementia in animal models and human studies ^18–26^.

Consistently, DNAm surrogates of smoking and of several plasma protein components of GrimAge2 were associated with cognitive aging in our sample of middle-aged and older Hispanic/Latino adults. The strongest associations across measures of cognitive aging and visits were for hemoglobin A1C. This is in line with our recent report on the association of elevated plasma levels of hemoglobin A1C with cognitive decline and risk of MCI in this cohort ^27^. It may also explain the slight outperformance of GrimAge2 over GrimAge in association analyses with cognitive aging in this sample.

Over 25 studies have investigated cross-sectional associations of one or more epigenetic clocks with measures of cognitive function in adulthood ^28,29^, although none in a large sample of Hispanic/Latino middle-aged and older individuals. While these studies differed in their measures of cognitive performance and epigenetic clocks assessed, with few exceptions ^30,31^, they reported a significant association of epigenetic aging with lower cognitive performance^15,17,32–40^. Our large sample size detected associations of all epigenetic clocks with global cognitive function at both visits, independent of education, language preference, and cardiovascular health. Consistent findings were observed for individual cognitive tests although associations of B-SEVLT tended to be weaker or non-significant with first-generation clocks.

In several studies, second- and third-generation clocks outperformed first-generation clocks in predicting cognitive performance ^15,17,37,40^. Similar to our findings, compared to other clocks, GrimAge acceleration showed the strongest associations with measures of cognitive function in 490 older adults from the Irish Longitudinal Study on Ageing ^40^, in 1115 middle-aged individuals from the Coronary Artery Risk Development in Young Adults (CARDIA) study ^17^, and in 3282 older adults from the Health and Retirement Study ^37^. Moreover, across various studies examining the association of epigenetic clocks with specific cognitive domains, the most consistent findings were for GrimAge acceleration associated with episodic memory and processing speed ^28^. In contrast, in 649 older adults from the Alzheimer’s Disease Neuroimaging Initiative (ADNI), DunedinPACE showed stronger and more consistent associations with multiple measures of cognitive function than other epigenetic clocks, including GrimAge ^15^.

Few studies have examined the association of epigenetic clocks with longitudinal decline in cognitive function in middle-aged or older adults ^17,31,41–43^. In all studies, there was an association of greater epigenetic age acceleration in at least one clock with faster cognitive decline over a follow-up period ranging from 5 to 15 years ^17,31,41–43^. Consistent with our findings, in studies examining multiple generation clocks, significant associations with longitudinal decline in cognitive function were generally observed with second- or third-generation clocks but not first-generation clocks ^17,43^.

The relationship between epigenetic clocks and MCI has not been widely studied. A study by Shadyab et al. found no association of accelerated epigenetic aging measured by first- and second-generation clocks with MCI in 578 elderly women (mean age 70 years) ^44^. However, in a subset of 262 women who developed coronary heart disease during follow-up, a higher Horvath age acceleration was associated with greater MCI risk ^44^. We observed significant associations between all clocks and MCI, with stronger associations identified for second- and third-generation clocks. Notably, associations of GrimAge acceleration with, both, MCI and presence of significant cognitive decline were observed with visit 1 measures, minimizing the possibility of reverse causation. Interestingly, with some exceptions ^15^, studies investigating the association of epigenetic aging with clinical dementia have largely yielded null results ^44–46^ but all these studies suffered from a limited sample size. More recently, in the large Framingham Heart Study Offspring Cohort (N=2264), there were significant associations of acceleration in the Horvath, PhenoAge, GrimAge, and DunedinPACE epigenetic clocks with incident dementia, with stronger effects observed for second- and third-generation clocks, notably DunedinPACE ^15^.

Only one study in 750 middle-aged non-Hispanic White and African American adults from the CARDIA cohort (mean age 50 years) has examined the association of longitudinal change in epigenetic clocks with measures of cognitive aging, and reported no association between the 5-year change in GrimAge acceleration with 5-year change in cognitive function ^17^. In contrast, we observed that a 7-year increase in age acceleration for second- and third-generation clocks was associated with significant cognitive decline over the same period and with a greater risk of MCI after accounting for visit 1 age acceleration, suggesting that longitudinal assessment of these clocks may provide additional value in predicting cognitive aging and impairment, beyond assessment at a single time point.

Age-related changes in DNA methylation caused by biological, environmental, and lifestyle factors are an important driver of brain aging and decline in brain and cognitive health ^47^. The mechanisms underlying the relationship between epigenetic age acceleration and cognitive aging are unknown but may reflect changes in metabolism, immunity, and autophagy ^48^, and perhaps other biological pathways that may overlap with those implicated in learning and memory. Epigenetic clocks have been shown to be heritable and genome-wide association studies have identified multiple genetic loci ^49^. Shared genetic variants between epigenetic aging and cognitive aging could also underlie the observed associations, as supported by significant genetic correlations of GrimAge and PhenoAge acceleration with cognitive traits ^49^. Despite considerable strengths, including a large sample of Hispanic/Latino adults and a 7-year longitudinal assessment of cognitive and epigenetic aging, our study suffers from several limitations. First, despite robust associations, the magnitude of effects of epigenetic clocks on measure of cognitive aging was generally modest, although that of change in clocks over time was considerably larger and similar to APOE4 effects. Second, while we adjusted for education, language preference, and cardiovascular health, residual confounding may remain. Third, only two time-points were available for both cognitive and epigenetic assessments, which may be more sensitive to random variation in the measurements rather than reflecting true trajectories. Additional data on a third visit will help address this concern. Finally, our findings necessitate replication in other cohorts of Hispanic/Latino adults.

In conclusion, acceleration in second- and third-generation epigenetic clocks was associated with decline in cognitive health and cognitive impairment in middle-aged and older Hispanic/Latino adults. In particular, GrimAge showed the strongest associations across measures of cognitive aging. With further validation, it may represent a valuable biomarker that may help identify individuals at risk of accelerated cognitive decline, who may benefit from early interventions to maintain or improve brain health. Repeated assessment of GrimAge may provide additional information about the continuous changes in the aging process and may be helpful for monitoring effectiveness of health-related interventions to improve cognitive health.

## Methods

### Study Participants

Participants were selected from the Study of Latinos Investigation of Neurocognitive Aging (SOL-INCA), an ancillary study of the Hispanic Community Health Study/ Study of Latinos (HCHS/SOL). The design, cohort selection and recruitment for HCHS/SOL and SOL-INCA have been previously described ^50–52^. Briefly, at the HCHS/SOL visit 1 (2008-2011), 16,415 Hispanic/Latino adults (ages 18-74) from six backgrounds (Central American, Cuban, Dominican, Mexican, Puerto Rican and South American) were enrolled from communities in the Bronx, NY; Chicago, IL; Miami, FL; and San Diego, CA. At visit 1, participants older than 45 years of age underwent a cognitive assessment and were re-assessed at visit 2 (2015-2018) as part of SOL-INCA (N=6377). SOL-INCA follows the complex study design of HCHS/SOL, including a multistage sampling strategy with stratification and clustering, and probability weights that account for non-response and attrition and permit valid inferences to targeted Latino populations in Bronx, Chicago, Miami, San Diego.^50,52^

For DNA methylation assessment, SOL-INCA participants with MCI and cognitively healthy controls from specified age and sex strata were selected (N=2800). The sampling strategy’s objective was to include all identified MCI cases identified in SOL-INCA and oversample cognitively normal participants from matching age and sex strata. The study was approved by the institutional review boards at each of the participating institutions. All human subjects provided written informed consent.

### Assessment of cognitive abilities and outcomes

Participants completed all assessments at each visit in their preferred language (Spanish or English). The neurocognitive tests administered at both visits were the Brief-Spanish English Verbal Learning Test B-SEVLT (sum and recall), which assesses verbal episodic learning and memory ^53^; the Word Fluency Test (WFT) of the Multilingual Aphasia Examination, which assesses verbal fluency ^54^; the Digit Symbol Substitution Test (DSST) of the Wechsler Adult Intelligence Scale-Revised, which assesses psychomotor speed and sustained attention ^55^; and the Six-Item Screener (SIS) derived from the Mini-Mental Status exam, which assesses mental status ^56^.

The primary outcomes of the study include a global cognitive function measure calculated at each visit as the average z-scores of B-SEVLT sum, B-SEVLT recall, WFT, and DSST scores; and the change in global cognition measure derived using a survey regression model to predict the cognitive score at visit 2 as a function of visit 1 cognitive score adjusting for the time elapsed between assessments. The global cognitive change value was calculated as (T2 − T2pred)/RMSE, where T2 represents the participant’s global cognitive function score at visit 2, T2pred is the predicted score, and RMSE is the root mean squared error of the fitted model. Details about this method have been published previously ^57^. Two binary cognitive outcomes assessed at visit 2 were also analyzed: presence of significant cognitive decline and MCI. Significant cognitive decline between the two visits was assessed based on a latent change score model that takes into account cognitive test scores ^52^. Presence of significant cognitive decline was defined as a change in global cognitive function score between the two visits exceeding -0.055 standard deviation (SD) per year. MCI was assessed according to the National Institute on Aging-Alzheimer’s Association (NIA-AA) criteria ^58^ as described previously ^52^. Briefly, a participant was classified with MCI if three conditions were satisfied: (1) any cognitive test score ≦ -1 standard deviation (SD) of the SOL-INCA robust norms adjusted for age, sex, education, and Picture Vocabulary Test scores, (2) a global cognition function decline by more than 0.055 SD per year between the visits, and (3) a participant self-reported cognitive decline based on an Everyday Cognition questionnaire ^52^.

### DNA Methylation Data

Genomic DNA was isolated from frozen peripheral blood leukocytes in the Advanced Research & Diagnostics Laboratory at the University of Minnesota using the FlexSTAR+ automated workflow (Autogen; Holliston, MA, USA) and provided to the Human Genetics Center Laboratory at the University of Texas Health Science Center at Houston for further processing. All sample handling used uniquely 2D barcoded Matrix vials (ThermoFisher Scientific; Waltham, MA, USA) integrated with the Human Genetics Center Laboratory Information Management System (LIMS). To minimize technical confounding and batch effects, samples were randomized to arrays so that visit 1 and visit 2 samples from a participant were placed on the same array and that there was approximately the same ratio of MCI cases to normal controls across arrays. Samples were randomly dispersed based on gender, Hispanic background, and field center. Approximately 500 ng of genomic DNA was bisulfite converted using the EZ-96 DNA Methylation Kit (Zymo Research Corporation; Irvine, CA, USA) and DNA methylation levels across ∼850,000 sites were measured using the Infinium MethylationEPIC v1.0 BeadChip (Illumina, Inc.; San Diego, CA, USA) using the manufacturer’s recommended protocols. Laboratory quality control included use of the BeadArray Controls Reporter (BACR) tool (Illumina, Inc.; San Diego, CA, USA) for review and exclusion of samples with poor bisulfite conversion efficiency, lower than normal staining, and poor hybridization. DNA methylation data were subjected to a comprehensive quality control (QC) protocol that uses both control-probe and data driven methods to filter out low-quality and questionable samples and probes. Probe signal detection and QC was performed using SeSaMe ^59^, which derives a p-value with out-of-band (OOB) array hybridization (pOOBAH) for signal detection for each probe and performs probe masking based on known EPIC array design issues as well as the signal detection p-value (Probes with pOOBAH<0.05 are masked). A total of 109,978 probes were masked after QC. Samples with sex mismatch, genotype mismatch, and those of poor quality (i.e., outliers in M-U plots, *outlyx*, and *bscon*)^60^ were excluded (N=65). A total of 2671 participants had DNA methylation data at both visits.

### Epigenetic Clocks Estimations

Principal component (PC)-trained clocks were estimated according to the method developed by Higgins-Chen et al.^61^ This method uses principal component analysis (PCA) to reduce variance in estimates of epigenetic age resulting from technical noise at individual CpG sites by leveraging multicollinearity in the data. At each visit, PC-trained DNA methylation age estimates were derived for four clocks: First-generation clocks, Horvath^7^ and Hannum^6^; and second-generation clocks, PhenoAge^8^ and GrimAge^9^. We also estimated a third-generation clock, DunedinPACE following the procedure described by Belsky et al.^5^. At each visit, estimates of age acceleration for each clock were calculated as the residuals from a linear regression of the DNA methylation age estimates on the chronological age. For each clock, change in age acceleration between the two visits was calculated as the difference in age acceleration between the two visits divided by the time (in years) between visits.

### Statistical Analysis

Descriptive statistics included pairwise Pearson’s r correlation coefficients were calculated to characterize the degree to which the various DNA methylation age estimates are related to one another and to chronological age at each visit, and across visits. Linear regression models were used to evaluate the associations of each measure of age acceleration with a global cognitive function score at each visit and with change in global cognitive function score between the two visits. Logistic regression models were used to evaluate the association of measures of age acceleration with presence of significant cognitive decline and with presence of MCI at V2. Model 1 was adjusted for age, sex, center and Hispanic background. Model 2 included additional adjustment for years of education and language preference. Model 3 also included adjustment for a measure of cardiovascular health (AHA Life’s Simple 7).^62^

## Data availability

The data used in this study are available from HCHS/SOL. Restrictions apply to the availability of these data, which were used under license for this study. Data are available from https://sites.cscc.unc.edu/hchs with the permission of HCHS/SOL.

## Code availability

Derivation of clocks used in this research was performed using codes available at: https://github.com/MorganLevineLab/PC-Clocks

https://github.com/danbelsky/DunedinPACE

https://github.com/perishky/meffonym

https://dnamage.genetics.ucla.edu/

## Acknowledgements and Funding

The authors thank the staff and participants of HCHS/SOL for their important contributions. A complete list of staff and investigators is available on the study website http://www.cscc.unc.edu/hchs/

This work was supported by grant RF1 AG061022. The Hispanic Community Health Study/Study of Latinos is a collaborative study supported by contracts from the National Heart, Lung, and Blood Institute (NHLBI) to the University of North Carolina (HHSN268201300001I / N01-HC-65233), University of Miami (HHSN268201300004I / N01-HC-65234), Albert Einstein College of Medicine (HHSN268201300002I / N01-HC-65235), University of Illinois at Chicago (HHSN268201300003I / N01-HC-65236 Northwestern Univ), and San Diego State University (HHSN268201300005I / N01-HC-65237). The following Institutes/Centers/Offices have contributed to the HCHS/SOL through a transfer of funds to the NHLBI: National Institute on Minority Health and Health Disparities,

National Institute on Deafness and Other Communication Disorders, National Institute of Dental and Craniofacial Research, National Institute of Diabetes and Digestive and Kidney Diseases, National Institute of Neurological Disorders and Stroke, NIH Institution-Office of Dietary Supplements.

## Authors’ Contributions

Study was conceptualized by M.F. and H.G. Data collection and analysis were performed by M.F., H.G, W.T., and R.X. The first draft of the manuscript was written by M.F., and all authors commented on different versions of the manuscript. All authors read and approved the final manuscript.

## Competing Interest

None

## Notes

### Competing Interest Statement

The authors have declared no competing interest.

### Author Declarations

The Institutional Review Board of the University of Texas Health Science Center at Houston gave…… ethical approval for this work.

