## Supplemental Tables for "Longitudinal associations of epigenetic aging with cognitive aging in Hispanic/Latino adults from the Hispanic Community Health Study/Study of Latinos"

**Supplementary Table 1.** Correlations between chronological age and DNA methylation age estimated from 5 epigenetic clocks at each visit.

|  | Hannum | Horvath | PhenoAge | GrimAge | DunedinPACE * |
| --- | --- | --- | --- | --- | --- |
| Visit 1 | 0.80 | 0.78 | 0.78 | 0.86 | 0.80 (0.05) |
| Visit 2 | 0.78 | 0.76 | 0.76 | 0.86 | 0.78 (0.13) |

P value < 0.0001 for all correlations; * rescaled values (original scale shown in parentheses)

**Supplementary Table 2.** Correlation between epigenetic age acceleration within visit and across visits. Pairwise correlations between clocks at visit 1 are shown in grey. Pairwise correlations between clocks at visit 2 are shown in white. Pairwise correlations between clocks across visits are shown in black.

|  | Hannum | Horvath | PhenoAge | GrimAge | DunedinPACE |
| --- | --- | --- | --- | --- | --- |
| Hannum | 0.89 | 0.88 | 0.68 | 0.34 | 0.21 |
| Horvath | 0.88 | 0.93 | 0.60 | 0.33 | 0.18 |
| PhenoAge | 0.70 | 0.64 | 0.81 | 0.50 | 0.45 |
| GrimAge | 0.41 | 0.38 | 0.57 | 0.91 | 0.53 |
| DunedinPACE | 0.27 | 0.21 | 0.48 | 0.53 | 0.81 |

P value < 0.0001 for all correlations;

**Supplementary Table 3.** Cross-sectional associations of epigenetic clocks with individual cognitive test scores at each visit.

Model 1: Adjusted for age, gender, center, and Hispanic background; Model 2: Adjusted for variables in Model 1 + years of education, language preference; Model 3: Adjusted for variables in Model 2 + cardiovascular health score (Life’s Simple 7 category); * DunedinPACE was rescaled to allow for comparison with other clocks (P-value shown for association on the original scale)

**3A.** B-SEVLT sum

|  |  | Model 1 | | | Model 2 | | | Model 3 | | |
| --- | --- | --- | --- | --- | --- | --- | --- | --- | --- | --- |
|  | **EAA Measure** | beta | SE | P value | beta | SE | P value | beta | SE | P value |
| Visit 1 | Hannum | -0.005 | 0.004 | 0.259 | -0.003 | 0.004 | 0.429 | -0.003 | 0.004 | 0.427 |
|  | Horvath | -0.007 | 0.004 | 0.089 | -0.005 | 0.004 | 0.155 | -0.005 | 0.004 | 0.180 |
|  | PhenoAge | -0.007 | 0.003 | **0.028** | -0.005 | 0.003 | 0.132 | -0.004 | 0.003 | 0.160 |
|  | GrimAge | -0.016 | 0.005 | **0.002** | -0.011 | 0.005 | **0.028** | -0.009 | 0.005 | 0.068 |
|  | DunedinPACE * | -0.010 | 0.002 | **5.4x10^-5^** | -0.007 | 0.002 | **0.002** | -0.006 | 0.002 | **0.009** |
| Visit 2 | Hannum | -0.012 | 0.005 | **0.008** | -0.010 | 0.004 | **0.019** | -0.009 | 0.004 | **0.036** |
|  | Horvath | -0.012 | 0.005 | **0.011** | -0.010 | 0.004 | **0.021** | -0.009 | 0.004 | **0.035** |
|  | PhenoAge | -0.016 | 0.003 | **6.3x10^-6^** | -0.015 | 0.003 | **2.5x10^-5^** | -0.013 | 0.003 | **0.0001** |
|  | GrimAge | -0.034 | 0.006 | **3.0x10^-8^** | -0.030 | 0.006 | **8.8x10^-7^** | -0.026 | 0.006 | **2.5x10^-5^** |
|  | DunedinPACE * | -0.014 | 0.003 | **6.8x10^-8^** | -0.011 | 0.003 | **7.4x10^-6^** | -0.009 | 0.003 | **0.0003** |

**3B.** B-SEVLT recall

|  |  | Model 1 | | | Model 2 | | | Model 3 | | |
| --- | --- | --- | --- | --- | --- | --- | --- | --- | --- | --- |
|  | **EAA Measure** | beta | SE | P value | beta | SE | P value | beta | SE | P value |
| Visit 1 | Hannum | -0.006 | 0.004 | 0.131 | -0.005 | 0.004 | 0.2121 | -0.005 | 0.004 | 0.252 |
|  | Horvath | -0.006 | 0.004 | 0.113 | -0.005 | 0.004 | 0.1763 | -0.005 | 0.004 | 0.249 |
|  | PhenoAge | -0.007 | 0.003 | **0.026** | -0.005 | 0.003 | 0.093 | -0.004 | 0.003 | 0.174 |
|  | GrimAge | -0.011 | 0.005 | **0.023** | -0.008 | 0.005 | 0.125 | -0.004 | 0.005 | 0.380 |
|  | DunedinPACE * | -0.008 | 0.002 | **0.0004** | -0.006 | 0.002 | **0.006** | -0.005 | 0.002 | **0.044** |
| Visit 2 | Hannum | -0.006 | 0.005 | 0.231 | -0.004 | 0.005 | 0.348 | -0.004 | 0.005 | 0.433 |
|  | Horvath | -0.004 | 0.005 | 0.374 | -0.003 | 0.005 | 0.505 | -0.003 | 0.005 | 0.563 |
|  | PhenoAge | -0.012 | 0.004 | **0.0008** | -0.011 | 0.003 | **0.002** | -0.010 | 0.004 | **0.005** |
|  | GrimAge | -0.020 | 0.006 | **0.0008** | -0.017 | 0.006 | **0.004** | -0.015 | 0.006 | **0.016** |
|  | DunedinPACE * | -0.008 | 0.003 | **0.003** | -0.006 | 0.003 | **0.029** | -0.004 | 0.003 | 0.106 |

**3C.** WF

|  |  | Model 1 | | | Model 2 | | | Model 3 | | |
| --- | --- | --- | --- | --- | --- | --- | --- | --- | --- | --- |
|  | **EAA Measure** | beta | SE | P value | beta | SE | P value | beta | SE | P value |
| Visit 1 | Hannum | -0.009 | 0.004 | **0.029** | -0.007 | 0.004 | 0.075 | -0.006 | 0.004 | 0.106 |
|  | Horvath | -0.011 | 0.004 | **0.012** | -0.008 | 0.004 | **0.027** | -0.008 | 0.004 | **0.043** |
|  | PhenoAge | -0.014 | 0.003 | **3.3x10^-5^** | -0.010 | 0.003 | **0.001** | -0.009 | 0.003 | **0.003** |
|  | GrimAge | -0.024 | 0.005 | **5.8x10^-6^** | -0.017 | 0.005 | **0.0005** | -0.016 | 0.005 | **0.002** |
|  | DunedinPACE * | -0.015 | 0.002 | **1.2x10^-9^** | -0.011 | 0.002 | **8.7x10^-7^** | -0.011 | 0.002 | **1.3x10^-5^** |
| Visit 2 | Hannum | -0.015 | 0.005 | **0.002** | -0.012 | 0.004 | **0.007** | -0.011 | 0.004 | **0.014** |
|  | Horvath | -0.015 | 0.005 | **0.001** | -0.013 | 0.004 | **0.003** | -0.012 | 0.004 | **0.005** |
|  | PhenoAge | -0.016 | 0.004 | **1.1x10^-5^** | -0.014 | 0.003 | **8.5x10^-5^** | -0.012 | 0.003 | **0.0003** |
|  | GrimAge | -0.033 | 0.006 | **1.4x10^-7^** | -0.025 | 0.006 | **6.3x10^-6^** | -0.023 | 0.006 | **0.0001** |
|  | DunedinPACE * | -0.016 | 0.003 | **4.3x10^-9^** | -0.011 | 0.002 | **3.2x10^-6^** | -0.010 | 0.003 | **0.0001** |

**3D.** DSST

|  |  | Model 1 | | | Model 2 | | | Model 3 | | |
| --- | --- | --- | --- | --- | --- | --- | --- | --- | --- | --- |
|  | **EAA Measure** | beta | SE | P value | beta | SE | P value | beta | SE | P value |
| Visit 1 | Hannum | -0.009 | 0.004 | **0.016** | -0.007 | 0.003 | **0.032** | -0.007 | 0.003 | **0.031** |
|  | Horvath | -0.011 | 0.004 | **0.005** | -0.008 | 0.003 | **0.010** | -0.009 | 0.003 | **0.008** |
|  | PhenoAge | -0.010 | 0.003 | **0.001** | -0.006 | 0.003 | **0.016** | -0.006 | 0.003 | **0.019** |
|  | GrimAge | -0.012 | 0.005 | **0.013** | -0.008 | 0.004 | **0.060** | -0.008 | 0.004 | **0.053** |
|  | DunedinPACE * | -0.010 | 0.002 | **2.8x10^-5^** | -0.006 | 0.002 | **0.003** | -0.006 | 0.002 | **0.002** |
| Visit 2 | Hannum | -0.012 | 0.004 | **0.003** | -0.010 | 0.004 | **0.006** | -0.009 | 0.004 | **0.011** |
|  | Horvath | -0.013 | 0.004 | **0.003** | -0.010 | 0.004 | **0.005** | -0.009 | 0.004 | **0.008** |
|  | PhenoAge | -0.012 | 0.003 | **0.0001** | -0.011 | 0.003 | **1.1x10^-4^** | -0.010 | 0.003 | **0.0004** |
|  | GrimAge | -0.027 | 0.005 | **5.9x10^-7^** | -0.023 | 0.005 | **3.8x10^-7^** | -0.022 | 0.005 | **2.4x10^-6^** |
|  | DunedinPACE * | -0.012 | 0.002 | **1.4x10^-7^** | -0.009 | 0.002 | **2.0x10^-5^** | -0.007 | 0.002 | **0.0002** |

**Supplementary Table 4.** Association of change in global cognitive function with epigenetic age acceleration (EAA) at visit 1 for 5 clocks

|  | Model 1 | | | Model 2 | | | Model 3 | | |
| --- | --- | --- | --- | --- | --- | --- | --- | --- | --- |
| **EAA Measure** | beta | SE | P value | beta | SE | P value | beta | SE | P value |
| V1 Hannum | -0.014 | 0.006 | **0.0177** | -0.013 | 0.006 | **0.026** | -0.012 | 0.006 | **0.032** |
| V1 Horvath | -0.010 | 0.006 | 0.076 | -0.009 | 0.006 | 0.106 | -0.009 | 0.006 | 0.112 |
| V1 PhenoAge | -0.015 | 0.005 | **0.0016** | -0.013 | 0.005 | **0.0040** | -0.012 | 0.005 | **0.008** |
| V1 GrimAge | -0.027 | 0.007 | **0.0002** | -0.026 | 0.007 | **0.0005** | -0.024 | 0.008 | **0.002** |
| V1 DunedinPACE * | -0.009 | 0.003 | **0.0028** | -0.008 | 0.003 | **0.0082** | -0.007 | 0.003 | **0.035** |
| V2 Hannum | -0.010 | 0.005 | 0.0653 | -0.009 | 0.005 | 0.0853 | -0.009 | 0.005 | 0.115 |
| V2 Horvath | -0.010 | 0.005 | 0.0639 | -0.009 | 0.005 | 0.0861 | -0.009 | 0.005 | 0.096 |
| V2 PhenoAge | -0.016 | 0.004 | **9.7x10^-5^** | -0.016 | 0.004 | **0.0002** | -0.015 | 0.004 | **0.0005** |
| V2 GrimAge | -0.032 | 0.007 | **9.2x10^-6^** | -0.031 | 0.007 | **1.7x10^-5^** | -0.029 | 0.007 | **0.0001** |
| V2 DunedinPACE * | -0.009 | 0.003 | **0.0006** | -0.009 | 0.003 | **0.0021** | -0.007 | 0.003 | **0.0112** |

Model 1: Adjusted for age, gender, center, and Hispanic background; Model 2: Adjusted for variables in Model 1 + years of education, language preference; Model 3: Adjusted for variables in Model 2 + cardiovascular health (Life’s Simple 7 category); * DunedinPACE was rescaled to allow for comparison with other clocks (P-value shown for association on the original scale)

**Supplementary Table 5.** Association of change in individual cognitive tests scores with change (Δ) in epigenetic age acceleration (EAA) between visit 1 and visit 2 for 5 clocks.

Model 1: Adjusted for V1 age acceleration, age, gender, center, and Hispanic background; Model 2: Adjusted for variables in Model 1 + years of education, language preference; Model 3: Adjusted for variables in Model 2 + cardiovascular health (Life’s Simple 7 category); * DunedinPACE was rescaled to allow for comparison with other clocks

**5A.** B-SEVLT sum change

|  | Model 1 | | | Model 2 | | | Model 3 | | |
| --- | --- | --- | --- | --- | --- | --- | --- | --- | --- |
| **EAA Measure** | beta | SE | P value | beta | SE | P value | beta | SE | P value |
| Δ Hannum | -0.028 | 0.061 | 0.642 | -0.025 | 0.060 | 0.681 | -0.015 | 0.061 | 0.811 |
| Δ Horvath | -0.090 | 0.076 | 0.240 | -0.090 | 0.076 | 0.237 | -0.079 | 0.076 | 0.301 |
| Δ PhenoAge | -0.075 | 0.038 | **0.048** | -0.078 | 0.038 | **0.039** | -0.069 | 0.038 | 0.070 |
| Δ GrimAge | -0.078 | 0.087 | 0.371 | -0.089 | 0.087 | 0.309 | -0.073 | 0.088 | 0.407 |
| Δ DunedinPACE * | -0.021 | 0.028 | 0.461 | -0.018 | 0.028 | 0.519 | -0.011 | 0.028 | 0.685 |

**5B.** B-SEVLT recall change

|  | Model 1 | | | Model 2 | | | Model 3 | | |
| --- | --- | --- | --- | --- | --- | --- | --- | --- | --- |
| **EAA Measure** | beta | SE | P value | beta | SE | P value | beta | SE | P value |
| Δ Hannum | 0.055 | 0.061 | 0.370 | 0.057 | 0.061 | 0.348 | 0.059 | 0.061 | 0.339 |
| Δ Horvath | 0.002 | 0.077 | 0.979 | 0.001 | 0.077 | 0.987 | -0.002 | 0.077 | 0.980 |
| Δ PhenoAge | -0.047 | 0.038 | 0.219 | -0.050 | 0.038 | 0.186 | -0.049 | 0.038 | 0.202 |
| Δ GrimAge | -0.089 | 0.089 | 0.314 | -0.100 | 0.088 | 0.258 | -0.093 | 0.089 | 0.293 |
| Δ DunedinPACE * | -0.010 | 0.028 | 0.722 | -0.008 | 0.028 | 0.783 | -0.005 | 0.029 | 0.857 |

**5C.** WF change

|  | Model 1 | | | Model 2 | | | Model 3 | | |
| --- | --- | --- | --- | --- | --- | --- | --- | --- | --- |
| **EAA Measure** | beta | SE | P value | beta | SE | P value | beta | SE | P value |
| Δ Hannum | 0.030 | 0.061 | 0.628 | 0.028 | 0.061 | 0.647 | 0.039 | 0.061 | 0.5198 |
| Δ Horvath | 0.005 | 0.077 | 0.950 | -0.002 | 0.076 | 0.980 | 0.007 | 0.077 | 0.9315 |
| Δ PhenoAge | -0.037 | 0.038 | 0.331 | -0.044 | 0.038 | 0.251 | -0.037 | 0.038 | 0.3284 |
| Δ GrimAge | -0.216 | 0.089 | **0.015** | -0.234 | 0.088 | **0.008** | -0.221 | 0.088 | **0.013** |
| Δ DunedinPACE * | -0.049 | 0.028 | 0.080 | -0.049 | 0.028 | 0.078 | -0.048 | 0.028 | 0.089 |

**5D.** DSST change

|  | Model 1 | | | Model 2 | | | Model 3 | | |
| --- | --- | --- | --- | --- | --- | --- | --- | --- | --- |
| **EAA Measure** | beta | SE | P value | beta | SE | P value | beta | SE | P value |
| Δ Hannum | -0.085 | 0.062 | 0.171 | -0.090 | 0.062 | 0.147 | -0.073 | 0.063 | 0.243 |
| Δ Horvath | -0.098 | 0.078 | 0.208 | -0.105 | 0.078 | 0.178 | -0.091 | 0.078 | 0.246 |
| Δ PhenoAge | -0.134 | 0.039 | **0.0006** | -0.139 | 0.039 | **0.0004** | -0.125 | 0.039 | **0.0015** |
| Δ GrimAge | -0.319 | 0.090 | **0.0004** | -0.334 | 0.090 | **0.0002** | -0.313 | 0.090 | **0.0005** |
| Δ DunedinPACE * | -0.058 | 0.029 | **0.042** | -0.059 | 0.029 | **0.040** | -0.051 | 0.029 | 0.079 |

**Supplementary Table 6.** Comparison of the magnitude of associations between change in DSST and epigenetic aging with the magnitude of association between change in DSST and APOE4

|  | beta | SE | P value |
| --- | --- | --- | --- |
| APOE4 alleles dosage | -0.084 | 0.042 | **0.043** |
| V1 PhenoAge | -0.010 | 0.004 | **0.019** |
| V1 GrimAge | -0.028 | 0.007 | **<0.0001** |
| V1 DunedinPACE* | -0.010 | 0.003 | **0.002** |
| Δ PhenoAge | -0.134 | 0.039 | **0.0006** |
| Δ GrimAge | -0.319 | 0.090 | **0.0004** |
| Δ DunedinPACE * | -0.058 | 0.029 | **0.042** |

Model adjusted for age, gender, center, and Hispanic background

**Supplementary Table 7.** Association of cognitive aging measures with estimates of GrimAge acceleration derived from multiple algorithms.

| **Cognitive aging measure** | **EAA Measure** | **Beta / Odds Ratio** | **SE / 95%CI** | **P value** |
| --- | --- | --- | --- | --- |
| V1 Global Cognitive Function | V1 PC GrimAge * | -0.018 | 0.004 | **9.1x10^-6^** |
|  | V1 GrimAge | -0.012 | 0.003 | **8.2x10^-5^** |
|  | V1 GrimAge2 | -0.012 | 0.003 | **9.9x10^-5^** |
| V2 Global Cognitive Function | V2 PC GrimAge * | -0.032 | 0.005 | **3.3x10^-10^** |
|  | V2 GrimAge | -0.020 | 0.004 | **8.4x10^-8^** |
|  | V2 GrimAge2 | -0.022 | 0.003 | **8.5x10^-11^** |
| MCI at V2 | V1 PC GrimAge * | 1.07 | 1.04; 1.11 | **5.4x10^-5^** |
|  | V1 GrimAge | 1.05 | 1.02; 1.07 | **6.6x10^-4^** |
|  | V1 GrimAge2 | 1.05 | 1.03; 1.08 | **2.5x10^-5^** |
|  | V2 PC GrimAge * | 1.09 | 1.05; 1.12 | **1.4x10^-6^** |
|  | V2 GrimAge | 1.06 | 1.04; 1.09 | **1.3x10^-6^** |
|  | V2 GrimAge2 | 1.06 | 1.04; 1.09 | **2.3x10^-7^** |
| Significant cognitive decline at V2 | V1 PC GrimAge * | 1.05 | 1.02; 1.08 | **1.6x10^-3^** |
|  | V1 GrimAge | 1.02 | 1.00; 1.05 | **2.9x10^-2^** |
|  | V1 GrimAge2 | 1.03 | 1.01; 1.05 | **4.5x10^-3^** |
|  | V2 PC GrimAge * | 1.07 | 1.04; 1.10 | **7.1x10^-6^** |
|  | V2 GrimAge | 1.05 | 1.03; 1.07 | **2.0x10^-5^** |
|  | V2 GrimAge2 | 1.05 | 1.03; 1.07 | **8.4x10^-7^** |

* Data shown in manuscript’s Tables. Models are adjusted for age, gender, center, and Hispanic background.

**Supplementary Table 8.** Association of DNA methylation components of GrimAge version 2 estimated at each visit with cognitive aging measures.

| **Visit 1** | V1 Global Cognitive Function | | MCI at V2 | | Significant cognitive decline at V2 | | Global Cognitive Function Change between V1 and V2 | |
| --- | --- | --- | --- | --- | --- | --- | --- | --- |
|  | Beta (SE) | P | Beta (SE) | P | Beta (SE) | P | Beta (SE) | P |
| ADM | -0.005 (0.013) | 0.69 | 0.165 (0.060) | **0.006** | 0.118 (0.048) | **0.0138** | -0.058 (0.024) | **0.0147** |
| B2M | -0.011 (0.011) | 0.33 | 0.106 (0.047) | **0.025** | 0.047 (0.040) | 0.24 | -0.036 (0.020) | **0.0694** |
| Cystatin C | -0.027 (0.011) | **0.015** | 0.115 ((0.048) | **0.017** | 0.087 (0.040) | **0.0303** | -0.053 (0.020) | **0.0081** |
| GDF15 | -0.015 (0.011) | 0.16 | 0.165 (0.045) | **0.0003** | 0.183 (0.042) | **1.4x10^-5^** | -0.070 (0.020) | **0.0004** |
| Leptin | -0.003 (0.017) | 0.87 | 0.038 (0.075) | 0.61 | 0.056 (0.062) | 0.36 | 0.012 (0.031) | 0.68 |
| logA1C | -0.034 (0.011) | **0.002** | 0.265 (0.048) | **3.3x10^-8^** | 0.232 (0.041) | **1.2x10^-8^** | -0.106 (0.020) | **8.0x10^-8^** |
| logCRP | -0.025 (0.011) | **0.030** | 0.141 (0.051) | **0.005** | 0.160 (0.042) | **0.0001** | -0.084 (0.020) | **3.9x10^-5^** |
| Pack Yrs | -0.042 (0.012) | **0.0003** | 0.167 (0.050) | **0.0007** | 0.104 (0.042) | **0.0144** | -0.052 (0.021) | **0.0139** |
| PAI1 | 0.007 (0.012) | 0.57 | 0.176 (0.052) | **0.0007** | 0.198 (0.043) | **3.8x10^-6^** | -0.074 (0.021) | **0.0004** |
| TIMP1 | -0.022 (0.011) | 0.05 | 0.098 (0.050) | 0.05 | 0.087 (0.041) | **0.0351** | -0.048 (0.020) | **0.0189** |

| **Visit 2** | V2 Global Cognitive Function | | MCI at V2 | | Significant cognitive decline at V2 | | Global Cognitive Function Change between V1 and V2 | |
| --- | --- | --- | --- | --- | --- | --- | --- | --- |
|  | Beta (SE) | P | Beta (SE) | P | Beta (SE) | P | Beta (SE) | P |
| ADM | -0.037 (0.017) | **0.0296** | 0.126 (0.061) | **0.0394** | 0.092 (0.049) | 0.06 | -0.052 (0.024) | **0.0341** |
| B2M | -0.028 (0.014) | **0.0414** | 0.062 (0.047) | 0.18 | 0.001 (0.040) | 0.98 | -0.010 (0.020) | 0.61 |
| Cystatin C | -0.042 (0.014) | **0.0023** | 0.042 (0.049) | 0.38 | 0.019 (0.040) | 0.63 | -0.016 (0.020) | 0.40 |
| GDF15 | -0.046 (0.014) | **0.0008** | 0.048 (0.046) | 0.31 | 0.035 (0.039) | 0.37 | -0.029 (0.019) | 0.13 |
| Leptin | -0.019 (0.021) | 0.36 | 0.027 (0.073) | 0.71 | 0.053 (0.060) | 0.38 | -0.005 (0.030) | 0.88 |
| logA1C | -0.077 (0.014) | **3.0x10^-8^** | 0.298 (0.047) | **3.1x10^-10^** | 0.187 (0.040) | **3.6x10^-6^** | -0.103 (0.020) | **1.7x10^-7^** |
| logCRP | -0.062 (0.015) | **2.2x10^-5^** | 0.173 (0.052) | **0.0008** | 0.116 (0.042) | **0.0059** | -0.075 (0.021) | **0.0003** |
| Pack Yrs | -0.054 (0.015) | **0.0002** | 0.128 (0.050) | **0.0106** | 0.059 (0.042) | 0.1594 | -0.042 (0.021) | **0.0447** |
| PAI1 | -0.038 (0.015) | **0.0117** | 0.196 (0.052) | **0.0002** | 0.126 (0.043) | **0.0032** | -0.075 (0.021) | **0.0004** |
| TIMP1 | -0.031 (0.014) | **0.0265** | 0.033 (0.049) | 0.50 | 0.016 (0.040) | 0.69 | -0.027 (0.020) | 0.17 |

Models are adjusted for age, gender, center, and Hispanic background.

ADM: Adrenomedullin; B2M: β2-microglobulin; GDF15: Growth differentiation factor 15; logA1c: hemoglobin A1C (log transformed); logCRP: C-reactive protein (log transformed); Pack Yrs: Smoking in pack/ year; PAI-1: Plasminogen activator inhibitor 1; TIMP1: tissue inhibitor metalloproteinase 1
